# Clinical characteristics and outcomes of critically ill patients with COVID-19 in a tertiary community hospital in upstate New York

**DOI:** 10.1101/2020.06.18.20135046

**Authors:** Jyotirmayee Lenka, Mamta S. Chhabria, Naman Sharma, Bryan E-Xin Tan, Leela Krishna Teja Boppana, Sharini Venugopal, Damanpaul S. Sondhi

**Affiliations:** Department of Medicine, Rochester General Hospital, Rochester, New York, USA; Department of Pulmonary and Critical Care Medicine, Rochester General Hospital, Rochester, New York, USA

**Keywords:** SARS-CoV-2, COVID-19, ICU, ventilator, New York

## Abstract

**Background:** There are limited reports describing critically ill COVID-19 patients in New York.

**Methods:** We conducted a retrospective analysis of 32 adult critically ill patients admitted to a tertiary community hospital in upstate NY, between March 14th and April 12th, 2020. We collected demographic, laboratory, ventilator, and treatment data, which were analyzed and clinical outcomes tabulated.

**Results:** 32 patients admitted to the ICU were included, with mean (±SD) follow-up duration 21 ± 7 days. Mean (±SD) age was 62.2 ± 11.2 years, and 62.5% were men. 27 (84.4%) of patients had one or more medical co-morbidities and 50% of the patients were current or former smokers. The mean (±SD) duration of symptoms was 6.6 (±4.4) days before presentation, with cough (81.3%), dyspnea (68.7%), and fever (65.6%) being most common. 23 (71.9%) patients received invasive mechanical ventilation. 5 (15.6%) had died, 11 (34.4%) had been discharged home, and 16 (50%) remained hospitalized, 8 (25%) of which were still in ICU. Mean (±SD) length of ICU stay was 10.2 (±7.7) days, and mean (±SD) length of hospital stay was 14.8 (±7.7) days.

**Conclusion:** Majority of patients were of older age and with medical co-morbidities. With adequate resource utilization, mortality of critically ill COVID-19 patients may not be as high as previously suggested.

## Main Body

## Introduction

Severe Acute Respiratory Syndrome Coronavirus 2 (SARS-CoV-2) is a novel coronavirus belonging to the *Coronaviridae* family, that has gripped the world in a pandemic of proportions last seen over a century ago during the Spanish flu outbreak of 1918. Since the coronavirus disease 2019 (COVID-19) was first identified in late December 2019 in Wuhan, China, there are now more than 2,980,000 confirmed cases globally, with the United States (US) leading the global tally with more than 980,000 cases. In the US, the state of New York (NY) has emerged as the epicenter of the outbreak with disproportionately large cases, totaling at 293,000 as of April 26^th^, 2020.

With high infectivity represented by an R0 of greater than 2^1^, human-to-human transmission and presence of a presymptomatic stage, SARS-CoV-2 led to an exponential growth of cases in a short period, overwhelming healthcare systems across the state and resulting in unprecedented effects on social, economic and healthcare sectors. This has galvanized hospital systems, including our own to come up with innovative means to handle the surge of cases at the peak of this pandemic, including expanding intensive care unit (ICU) teams, personal protective equipment conservation strategies, and grim conversations about resource allocation.

SARS-Cov-2 is an enveloped virus with a large plus-strand RNA genome, and acts primarily as a respiratory pathogen, infecting cells by attaching to ACE-2 receptors. The clinical spectrum of COVID-19 ranges from mild to critically ill cases^2^ with reports indicating 5-9% of all cases are admitted to the ICU with severe respiratory failure^3, 4^.

Despite several observational studies and case series on COVID-19 patients from the inpatient and outpatient setting, there are currently limited reports describing critically ill patients in the US.^5, 6^ In this case series, we describe demographic characteristics, co-existing conditions, ventilation parameters, and clinical outcomes of patients admitted to the medical ICU at Rochester General Hospital (RGH), a tertiary community hospital in Monroe county, NY, which also functions as a safety net hospital for the area. We aim to help guide identification of those at greatest risk of deterioration, and improve decision making in managing this unique subset of patients.

## Methods

### Study population and institutional approval

We included adult patients, 18 years or older, with laboratory-confirmed COVID-19 infection who were admitted to the medical intensive care unit (ICU) at RGH or transferred to RGH from other community hospitals between March 14^th^ and April 12^th^, 2020. These were then followed up until April 18, 2020. We excluded pregnant or incarcerated patients, patients aged < 18 years of age, and patients requiring less than 6 L of supplemental oxygen. Rochester Regional Health (RRH) Institutional Review Board approved our case series (IRB:1982A), informed consent was waived and researchers analyzed only de-identified data.

### Data collection

We collected demographic, clinical, laboratory, radiological, ventilator and treatment data by manual review of electronic medical records (EPIC). These were then analyzed to tabulate clinical outcomes. All documentation, investigations, and management of patients, had been performed at the discretion of the primary treatment team. A laboratory-confirmed case of COVID-19 was defined as a positive result on the SARS-CoV-2 real-time reverse transcriptase-polymerase-chain-reaction (RT-PCR) assay of nasopharyngeal or oropharyngeal swab or lower respiratory tract specimens. Specimens were obtained and processed according to CDC guidelines.^7^ Until April 10^th^ we utilized RRH laboratory-developed manual PCR assay with emergency use authorization from the CDC for inpatient use and kits from CDC at public health laboratories Buffalo or Wadsworth for outpatient use. After April 10^th^, we used the Cobas ® 6800 System by Roche for inpatient and out-patient tests.

### Statistical Analysis

We present categorical variables as counts and percentages. We present continuous variables as mean ± standard deviation (SD) or median and interquartile range (IQR), wherever appropriate. Data were analyzed using the following statistical tests: independent sample t-test, Wilcoxon-Mann Whitney test, Fisher Exact test, and Chi-square, as appropriate. The analysis was performed using SPSS Statistics for Windows, version 24.0 (IBM Corp., Armonk, N.Y., USA). Two-tailed P values of < 0.05 were deemed statistically significant.

## Results

### Demographics and Presenting features: (Table 1)

We identified 32 critically ill patients admitted to the RGH ICU between March 14, 2020 and April 12, 2020. The mean (±SD) follow-up duration was 21 days (±7 days), with a minimum of 7 and a maximum of 35 days. Demographic characteristics of these patients are detailed in Table 1. Mean (±SD) age was 62.2 ±11.2 years, and 62.5% were men. 27 (84.4%) of patients had one or more medical comorbidities, of which obesity (68.8%) and hypertension (65.6%) were the most prevalent. 50% of them were either current or former smokers.

**Table 1:** Baseline characteristics and presenting symptoms

| Characteristics | Patients (N=32) |
| --- | --- |
| Age – years (Mean, ±SD, range) | 62.2 ± 11.2 (37-84) |
| Male | 20 (62.5%) |
| Female | 12 (37.5%) |
| **Ethnicity:** number (percent) |  |
| Hispanic | 12 (37.5%) |
| African American | 10 (31.2%) |
| Caucasian | 8 (25%) |
| Asian | 2 (6.25%) |
| **Comorbidities:** number (percent) |  |
| Coronary artery disease/heart failure | 14 (43.7%) |
| Hypertension | 21 (65.6%) |
| COPD/asthma | 7 (21.9%) |
| Diabetes Mellitus | 15 (46.9%) |
| Chronic Kidney Disease | 5 (15.6%) |
| BMI>30 | 22 (68.8%) |
| Smoking History | 16 (50%) |
| **Presenting Complaints:** number |  |
| (percent) |  |
| Fever | 21 (65.6%) |
| Cough | 26 (81.3%) |
| Sputum production | 14 (43.8%) |
| Dyspnea | 22 (68.7%) |
| Sore throat | 1 (3.1%) |
| Headache | 7 (21.9%) |
| Fatigue | 22 (68.8%) |
| Myalgia | 14 (43.8%) |
| Syncope/pre-syncope | 5 (15.6%) |
| Diarrhea/GI symptoms | 13 (40.6%) |
| Chills | 18 (56.2%) |
| Altered mental status | 5 (15.6%) |
| **Duration of symptoms - days** Mean,<br>(±SD), {range} | 6.6 (±4.4) {1-21} |

The mean (±SD) duration of symptoms was 6.6 (±4.4) days before presentation, with cough (81.3%), dyspnea (68.7%), and fever (65.6%) being most common. Other symptoms included diarrhea, fatigue, and myalgia. The median (IQR) temperature on presentation was 102.2 degrees Fahrenheit (99.8 - 103.1); median (IQR) oxygen saturation by pulse oximetry was 89% (82-93%). 62.5% patients were hypoxic and 35% of these were hypoxic without reported dyspnea.

### Laboratory, Radiology and Microbiological findings: (Table 2)

Table 2 shows both the initial and extreme laboratory values on ICU admission and during hospital stay. Lymphopenia was common, with median (IQR) Absolute lymphocyte count (ALC) of 1.1 (0.6 - 1.4) × 10^3^ cells per cubic millimeter, and a third of the cases (34.4%) developed severe lymphopenia (with ALC ≤ 0.3 ×10^3^ cells per cubic millimeter) during their ICU stay. The median (IQR) C-Reactive Protein (CRP) was 137.4 (78.1 - 181.8) mg/L and majority (68.75%) had initial CRP of ≥100 mg/L. Initial procalcitonin value of ≥0.5 was noted in 10 (31.25%) of the 32 patients, 5 of which had a positive sputum culture. D-dimer was elevated with a median initial D-dimer of 693 ng/mL, reaching extremely high values (>7650 ng/mL) in 11 (34.4%) of the 32 patients during hospital stay. Initial chest X-ray was abnormal in 22 (66.6 %) patients. A CT chest was obtained in 6 patients, all of whom had abnormal findings.

**Table 2:**
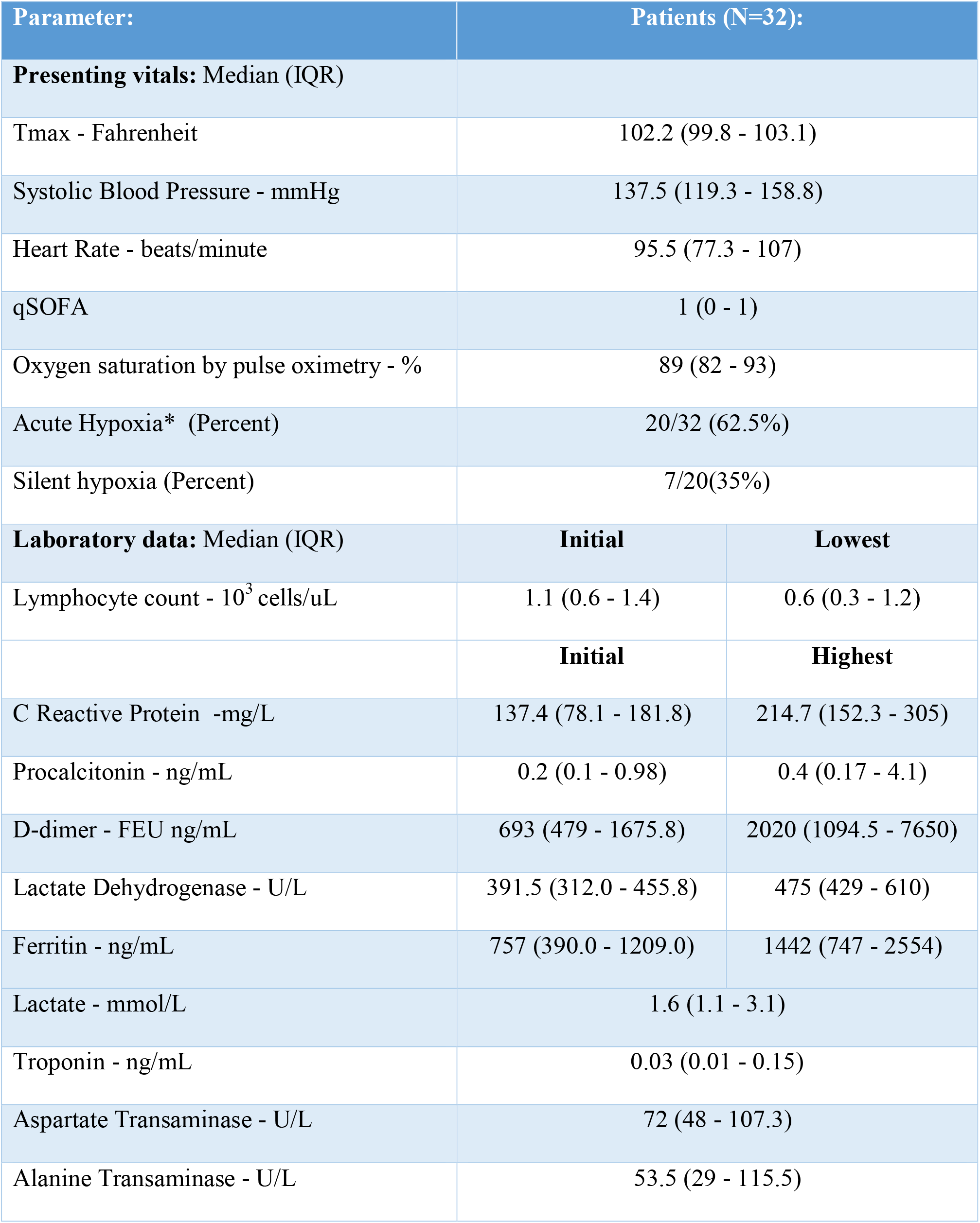
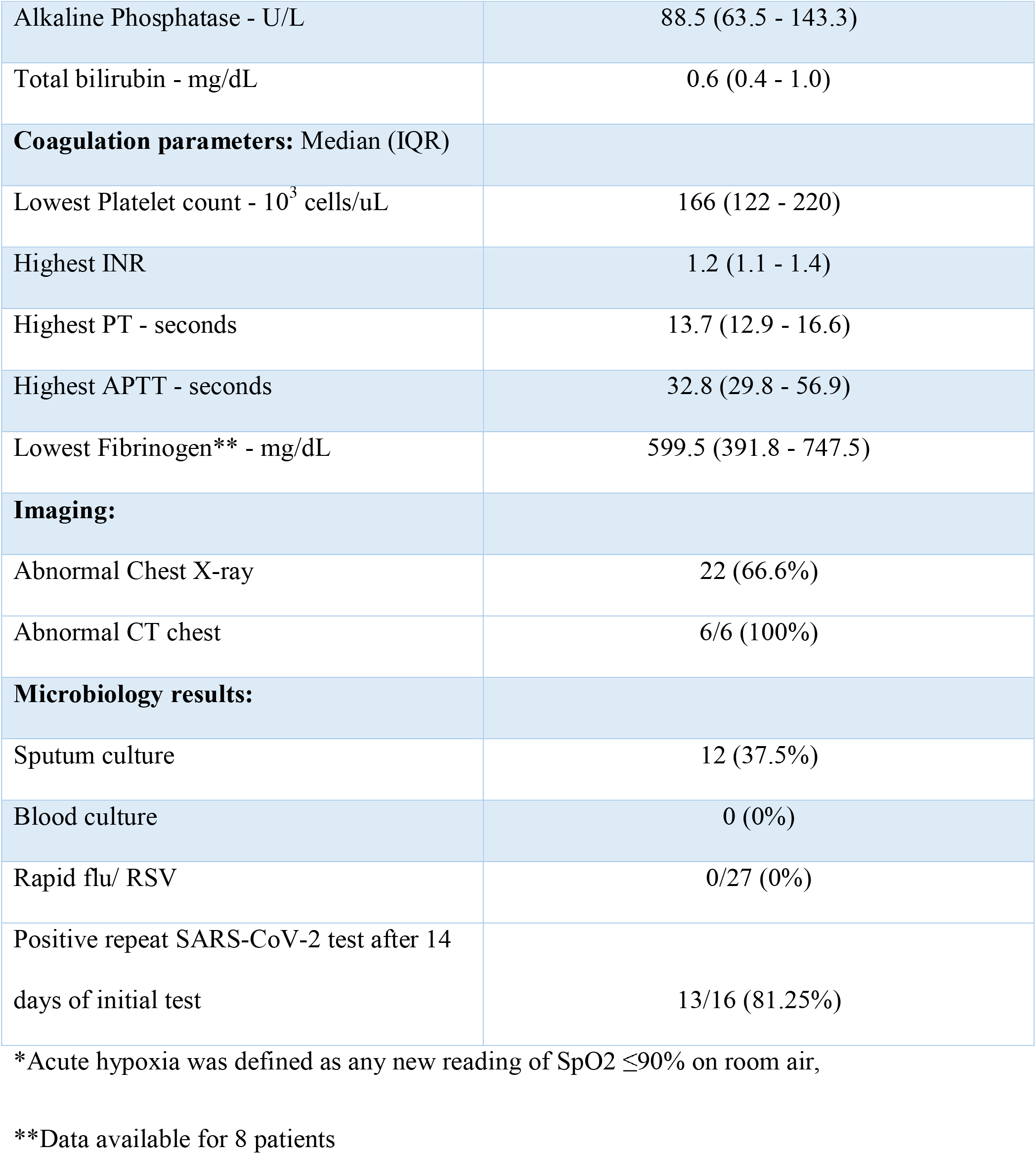
Presenting vitals, Laboratory data, Imaging findings and Microbiology data

| Parameter: | Patients (N=32): |  |
| --- | --- | --- |
| **Presenting vitals: Median (IQR)** |  |  |
| Tmax - Fahrenheit | 102.2 (99.8 - 103.1) |  |
| Systolic Blood Pressure - mmHg | 137.5 (119.3 - 158.8) |  |
| Heart Rate - beats/minute | 95.5 (77.3 - 107) |  |
| qSOFA | 1 (0 - 1) |  |
| Oxygen saturation by pulse oximetry - % | 89 (82 - 93) |  |
| Acute Hypoxia* (Percent) | 20/32 (62.5%) |  |
| Silent hypoxia (Percent) | 7/20(35%) |  |
| **Laboratory data: Median (IQR)** | **Initial** | **Lowest** |
| Lymphocyte count - 10<sup>3</sup> cells/uL | 1.1 (0.6 - 1.4) | 0.6 (0.3 - 1.2) |
|  | **Initial** | **Highest** |
| C Reactive Protein -mg/L | 137.4 (78.1 - 181.8) | 214.7 (152.3 - 305) |
| Procalcitonin - ng/mL | 0.2 (0.1 - 0.98) | 0.4 (0.17 - 4.1) |
| D-dimer - FEU ng/mL | 693 (479 - 1675.8) | 2020 (1094.5 - 7650) |
| Lactate Dehydrogenase - U/L | 391.5 (312.0 - 455.8) | 475 (429 - 610) |
| Ferritin - ng/mL | 757 (390.0 - 1209.0) | 1442 (747 - 2554) |
| Lactate - mmol/L | 1.6 (1.1 - 3.1) |  |
| Troponin - ng/mL | 0.03 (0.01 - 0.15) |  |
| Aspartate Transaminase - U/L | 72 (48 - 107.3) |  |
| Alanine Transaminase - U/L | 53.5 (29 - 115.5) |  |

|  |  |
| --- | --- |
| Alkaline Phosphatase - U/L | 88.5 (63.5 - 143.3) |
| Total bilirubin - mg/dL | 0.6 (0.4 - 1.0) |
| **Coagulation parameters: Median (IQR)** |  |
| Lowest Platelet count - $10^3$ cells/uL | 166 (122 - 220) |
| Highest INR | 1.2 (1.1 - 1.4) |
| Highest PT - seconds | 13.7 (12.9 - 16.6) |
| Highest APTT - seconds | 32.8 (29.8 - 56.9) |
| Lowest Fibrinogen** - mg/dL | 599.5 (391.8 - 747.5) |
| **Imaging:** |  |
| Abnormal Chest X-ray | 22 (66.6%) |
| Abnormal CT chest | 6/6 (100%) |
| **Microbiology results:** |  |
| Sputum culture | 12 (37.5%) |
| Blood culture | 0 (0%) |
| Rapid flu/ RSV | 0/27 (0%) |
| Positive repeat SARS-CoV-2 test after 14 days of initial test | 13/16 (81.25%) |

\*Acute hypoxia was defined as any new reading of SpO2 ≤ 90% on room air,
\*\*Data available for 8 patients

All patients had laboratory-confirmed testing of COVID-19; of these 27 were also tested for Influenza A, Influenza B and Respiratory Syncytial Virus, all of which were negative for co-infections with these respiratory viruses. 16 of the 32 patients had follow up repeat SARS-CoV-2 RT PCR testing after 14 days of their initial test, of which 13 (81.25%) resulted positive.

### ICU treatment characteristics: (Table 3)

23 (71.9%) of the patients received intubation and invasive mechanical ventilation (IMV) and the remaining 9 were managed with noninvasive ventilatory (NIV) support of which 4 received supplemental oxygen through high flow nasal cannula (HFNC). At that time, we did not use other modes of positive pressure ventilation like bilevel positive airway pressure (BiPAP) or continuous positive airway pressure (CPAP) in COVID-19 patients due to concerns of aerosolization.

**Table 3:** Ventilatory data, other ICU therapies and Medications used

| Parameter | Patient (N=32<sup>^</sup>) |
| --- | --- |
| **Respiratory support:** |  |
| High Flow Nasal Cannula | 5 (15.6%) |
| Intermediate Flow Nasal Cannula | 4 (12.5%) |
| Invasive Mechanical Ventilation (IMV) | 23 (71.9%) |
| **Characteristics of IMV: Median (IQR)** |  |
| Lowest pre-intubation PaO<sub>2</sub>/FiO<sub>2</sub> ratio * | 101 (75.8 - 183.8) |
| FiO<sub>2</sub> 4 hrs after intubation - percentage of oxygen | 70 (50 - 80) |
| PEEP max - cmH<sub>2</sub>O | 16 (14 - 20) |
| Plateau pressure 4 hrs after intubation- cmH<sub>2</sub>O | 26 (23 - 30.5) |
| Positive Inspiratory Pressure max- cmH<sub>2</sub>O | 29 (28 - 35) |
| Static compliance 4 hrs after intubation- cmH<sub>2</sub>O ** | 44 (31 - 59) |
| Driving pressures- cmH<sub>2</sub>O | 12 (0 – 14) |
| **ICU therapies:** |  |
| Intravenous vasopressors | 12/32 (36.3%) |
| Neuromuscular blockade | 9/23 (39.1%) |
| Intravenous pulmonary vasodilators | 8/23 (34.8%) |
| Prone positioning | 7/23 (30.4%) |
| Extracorporeal membrane oxygenation (ECMO) | 3/23 (13.0%) |
| Peri-intubation intravenous fluids | 15/23 (65.2%) |
| **Medication details:** |  |
| Hydroxychloroquine | 19 (59.4%) |
| Lopinavir-Ritonavir | 5 (15.6%) |
| Remdesivir | 0 (0%) |
| Tocilizumab | 3 (9.38%) |
| Azithromycin | 19 (59.4%) |
| Systemic Glucocorticoids | 15 (46.9%) |
| Statins | 19 (59.4%) |
| Concurrent antibiotics | 26 (8.1%) |
| ACE-i/ARB prior to hospitalization or during hospitalization | 10 (31.3%) |
| Thromboprophylaxis | 32 (100%) |

^N=32 unless specified, \*data available for 28 patients; we estimated PaO<sub>2</sub> from SpO<sub>2</sub> in 4 patients who did not have pre-ventilation arterial blood gas values available. \*\*data available for 11 out of 23 patients intubated. ACE-I = angiotensin converting enzyme inhibitor, ARB = angiotensin receptor blocker

For the 28 patients with data available, the median (IQR) ratio of the partial pressure of arterial oxygen to the fraction of inspired oxygen (PaO2/FiO2) ratio was 101 (75.8 - 183.8) consistent with moderate to severe Acute Respiratory Distress Syndrome (ARDS); median (IQR) FiO2 of 70% (50-80%) at 4 hours post-intubation. Among the 23 patients who received IMV, Pressure Control Ventilation (PCV) was the most common ventilator mode selected (in 52.2%), mostly based on the preference of treating intensivist. Median (IQR) positive end expiratory pressure (PEEP) used was 16 cmH_2_O (14-20 cmH_2_O) and median (IQR) plateau pressure was relatively high at 26 cmH_2_O (23 - 30.5) cm H_2_O. Median (IQR) static lung compliance was 44 (31 - 59) cm H_2_O in 11 patients with data available, none were less than 10 cmH_2_O. Median (IQR) driving pressure was 12 cmH_2_O (0 – 14) cmH_2_O. 7 of the 23 patients (30.4%) underwent prone positioning.

Of the 32, approximately one third (34.8%) received intravenous pulmonary vasodilators (epoprostenol). 3 patients required extra corporeal membrane oxygenation (ECMO), 2 veno-venous and 1 veno-arterial). 12 (36.3%) required vasopressor support.

19 (59.4%) of the patients received Hydroxychloroquine and the same number received Azithromycin and statins during hospitalization. 5(15.6%) received lopinavir-ritonavir, while 3 (9.38%) received tocilizumab. None of the patients received Remdesivir. Just under a half (46.9%) received steroids during ICU stay. A third (31.3%) were on ACE-I/ARBs prior to admission.

### Clinical Outcomes [Figure 1, Table 4 (4a, 4b)]

Follow up was complete until April 18, 2020, with mean (±SD) follow-up of 21 (±7) days, minimum of 7 days and a maximum of 35 days. Figure 1 outlines the disposition status of all 32 patients and Table 4 (4a, 4b) list our clinical outcomes. Of the 32 patients, 5 (15.6%) had died, 11 (34.4%) had been discharged home, and 16 (50%) remained hospitalized, 8 (25%) of which were still in ICU. A greater percentage of patients aged 60 years or above had died compared to patients under 60 years (21% vs. 7.7%). Of the 23 patients that received IMV, 11 (47.8%) had been extubated during follow up, with mean (±SD) number of days of invasive mechanical ventilation of 10 (±6.85) days. 8 remained intubated, with mean (±SD) ventilator days of 14.9 (± 6.7) days. The mean (±SD) length of ICU stay among all patients was 10.2 (±7.7) days, and mean (±SD) length of hospital stay was 14.8 (±7.7) days. In those still hospitalized, mean (±SD) length of stay was 18 (±7.4) days which is likely to be an underestimate. The lengths of ICU stay and hospital stay were significantly higher in those who received IMV than those managed by NIV (12.8 vs 3.4 days and 16.9 vs 9.2 days) respectively.

**Figure 1:**
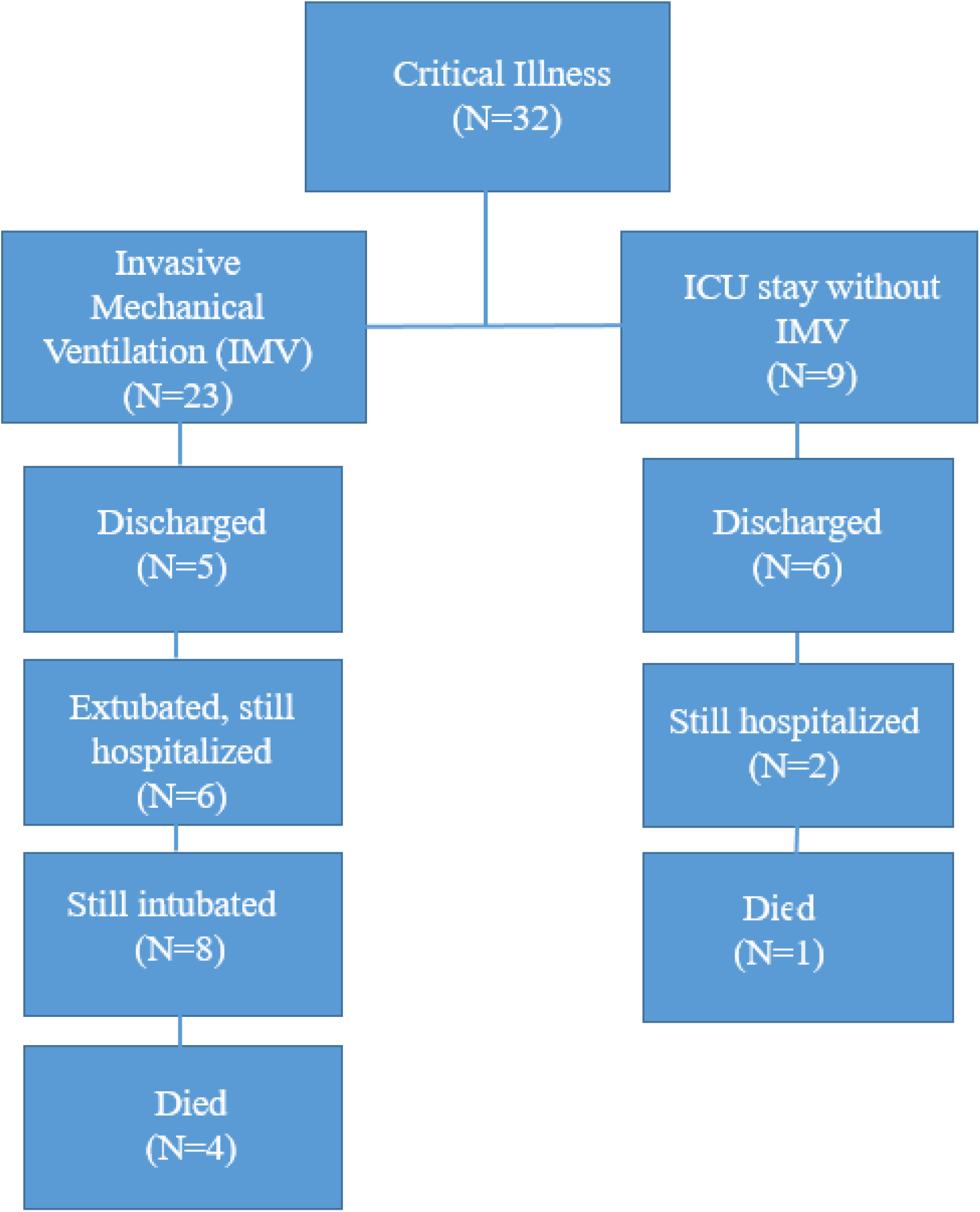
Disposition of patients at the time of analysis of results:

**Table 4a:** Clinical outcomes, as per status of invasive mechanical ventilation

| Clinical Outcome: Mean (±SD) | All patients: | IMV (N=23) | NIV (N=9) | P value |
| --- | --- | --- | --- | --- |
| Days on ventilator for those extubated; n=11^ | 10(6.8) |  |  |  |
| Days on ventilator, for those still intubated; n=8 | 14.9 (6.7) |  |  |  |
| Length of ICU stay -- days | 10.2 (7.7) | 12.8 (7.4) | 3.4 (3.1) | **<0.001** |
| Length of hospital stay -- days | 14.8 (7.7) | 16.9 (7.9) | 9.2 (2.9) | **0.006** |
| Length of hospitalization in those still hospitalized -- days | 18 (7.4) | 20.1 (6.9) | 10.5 (1.7) | **0.009** |
| Number discharged from ICU (excluding mortality); N=32 (percent) | 16 (32%) | 9 (39.1%) | 7 (77.8%) | 0.12 |
| Number discharged from hospital (excluding mortality); N=32 (percent) | 11 (34.4%) | 5 (21.7%) | 6 (66.7%) | 0.046 |
| Death; N=32 ( percent) | 5 (15.6%) | 4 (17.4%) | 1 (11.1%) | 1 |
| Duration of follow up -- days | 20.8 (7.4) |  |  |  |

^this excludes 1 patient who underwent compassionate extubation, and 3 deaths

**Table 4b:**
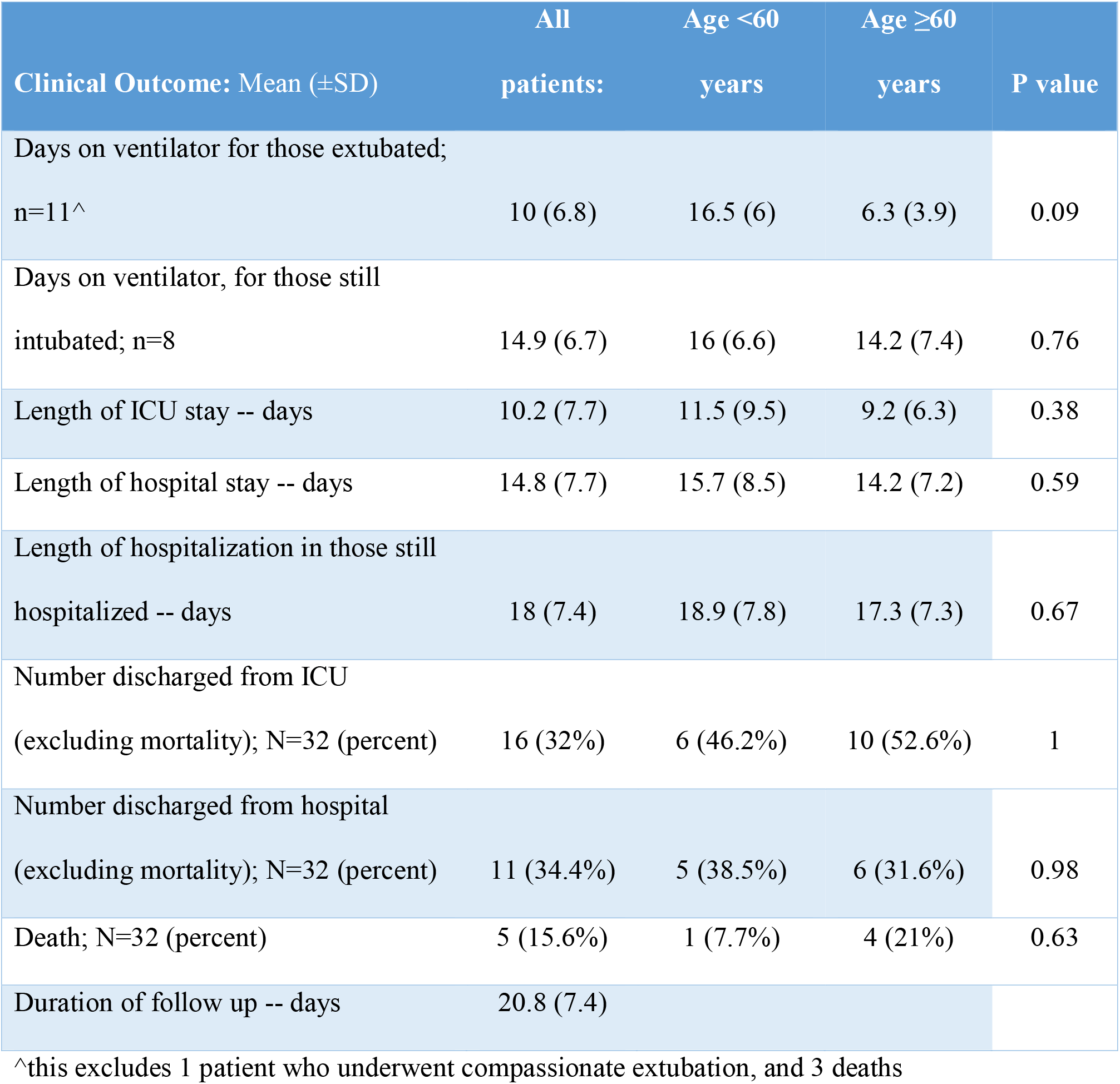
Clinical outcomes, as per age < or ≥60 years

### Complications

5 (15.6%) patients had adverse venous thrombotic-embolic events (VTE) with 1 patient (3%) developing massive pulmonary embolism (PE), 2 patients (6.2%) developing deep vein thrombosis (DVT)-1 patient (3%) developing extensive clots in hemodialysis catheter requiring therapeutic anticoagulation, and 1 patient (3%) developed bilateral embolic strokes. All were on pharmacological VTE prophylaxis with weight-based heparin, or low molecular weight heparin (LMWH). 2 patients (6.2%) developed renal failure necessitating renal replacement therapy (RRT) and 1 patient (3%) developed an ST-segment elevation Myocardial infarction (STEMI) requiring coronary artery bypass graft (CABG) and subsequent VA-ECMO support. Transaminitis and myocarditis were uncommon.

## Discussion

To our knowledge, this is the first case series exclusively on critically ill COVID-19 patients in the state of NY. Older persons, men and those with medical comorbidities were common in our series, suggesting that these patients may be at higher risk of severe illness and ICU admission, findings similar to other reports from New York City and Seattle^7, 5^. Fever, cough and dyspnea seem to be the most common symptoms. Since a majority of the patients had CRP ≥100, this could be used in creating risk calculators to identify patients at higher risk of critical illness. Interestingly only half of our cases with elevated procalcitonin had bacterial infections, suggesting that procalcitonin is more likely a marker of sepsis in the critically ill than of bacterial infection, corresponding to prior published studies^8^. Initial chest X-ray was normal in a third (33.4%) of patients, therefore a normal X-ray cannot rule out infection with COVID-19.

Secondary infection with positive sputum cultures were noted in a third (37.5%) of the patients, incidence similar to that in seasonal influenza^9^. Yet, co-infections with other respiratory viruses were not noted. As the protocol developed in our hospital for repeat testing of SARS-CoV-2, we noted that a very large number (13/16, 81.3%) of the patients tested positive after 2 weeks. Whether this implies continued infectivity, viral shedding, or mere inert viral RNA remains unclear.

Profound hypoxia seems to be the driving factor leading to intubation, as majority (62.5%) of the patients were hypoxic; interestingly, about a third [7/20, (35%)] of these were ‘silently’ hypoxic, with no reported dyspnea. To explain this Conde et al, describe an interesting theory of the virus manifesting neurotropism^10^, by involving the midbrain, respiratory and cardiovascular control centers, leading to decreased perception of dyspnea, despite hypoxia.

We noted low pre-ventilation PaO2/FiO2 ratios, consistent with ARDS definition by the Berlin criteria.^11^ However, our median driving pressures and static compliance values suggest findings of near-normal compliance, which is unusual for ARDS. Higher PEEP was used early on but with more experience, intensivists started using lower PEEP. Prone positioning was attempted in a third of patients receiving IMV, to help improve oxygenation by increasing alveolar recruitment^12^. The threshold to prone was relaxed from strict PROSEVA study^13^ indications to anyone with refractory hypoxemia; 2 patients on HFNC were tried on awake prone positioning, with improved oxygenation. The median FiO2 needs were high, with most patients requiring 50-60% FiO2 for long periods of time, underscoring prolonged profound hypoxia in this illness. Most patients were able to draw good tidal volumes despite low inspiratory pressures. These findings raise the question about the factors involved in pathogenesis and response to treatment in COVID-19. The findings of severe hypoxemia with preserved compliance have been noted in recent literature^14^. Current evidence on the pathophysiology of hypoxia in COVID-19 is changing. The hypoxia is hypothesized to be from 3 mechanisms, dysregulation of pulmonary perfusion, pulmonary microthrombi and ARDS.^15^ Gatinnoni et al^16^ recently described two possible phenotypes of patients with respiratory failure from COVID-19; Type L -characterized by near normal pulmonary compliance and low recruitability, low PEEP response wherein the primary pathology is theorized to be loss of hypoxic vasoconstriction and possibly pulmonary microthrombi; Type H on the other hand behaves more like traditional ARDS with poor pulmonary compliance and hence better response to high PEEP ventilation and lung recruitment techniques. Type L is peculiar for hypoxia out of proportion to lung infiltrates on imaging.^16^ As none of our deceased patients had autopsies (these being cancelled due to high risk of exposure), it is difficult for us to associate our findings with these theories.^17^ Due to such variability in response, personalized ventilator settings and management have been advised for COVID-19 patients.^18^ Figure 2 demonstrates the proposed ventilatory strategies for management of critically ill COVID-19 patients at RGH.

**Figure 2:**
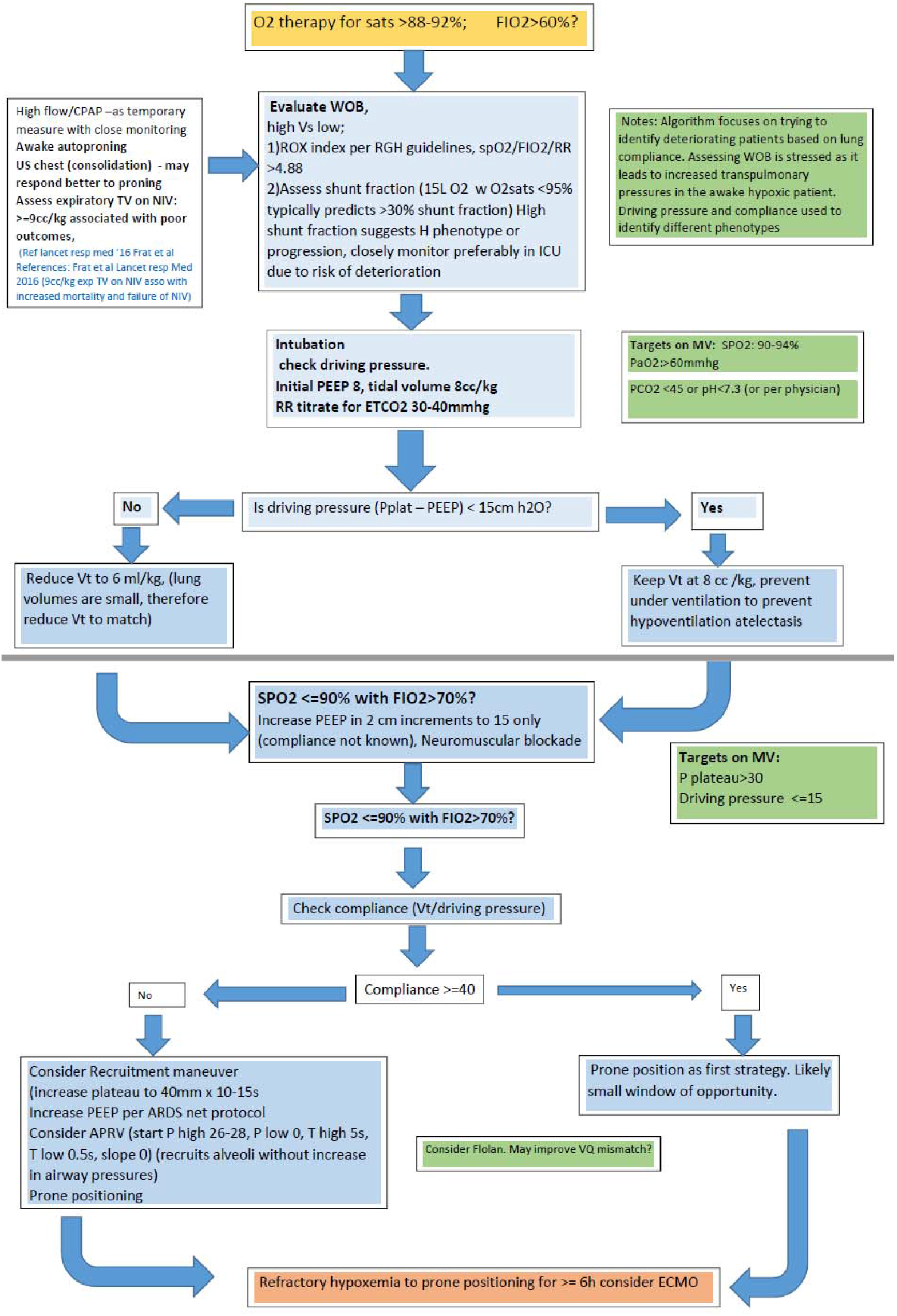
Proposed ventilation strategies for management of critically ill COVID-19 patients at RGH. ^inspired by ESICM webinar on ventilation strategies in COVID-19^15^

Among other ICU therapies, epoprostenol was used in a third of the patients, and a variable response was perceived. It has been noted that in some cases of COVID-19, despite low PaO2/FiO2 ratio, perfusion is maintained, which along with atelectasis, leads to a right to left shunt phenomenon^19^. Therefore pulmonary vasodilators such as epoprostenol may not be useful in these cases. Only a third of the patients required vasopressor support, and shock and multiorgan failure were uncommon in our patient series. These findings are inconsistent with some prior reports.^5^

The choice of COVID19-directed medication therapy was based on judgment of the infectious disease specialists and treating intensivists, and mostly based on an indigenous institutional algorithm [Figure 3 (3a, 3b)]. Just above half the patients received Hydroxychloroquine or Azithromycin during their hospitalization. None of the patients received Remdesivir. When compared to the recent study of compassionate use of Remdesivir by Grien et al^20^, our patient population appeared to have had similar baseline characteristics and outcomes in terms of mortality, rate of extubation, ICU LOS and hospital discharges. Yet, we have insufficient data to draw meaningful associations about these medications.

**Figure 3a:**
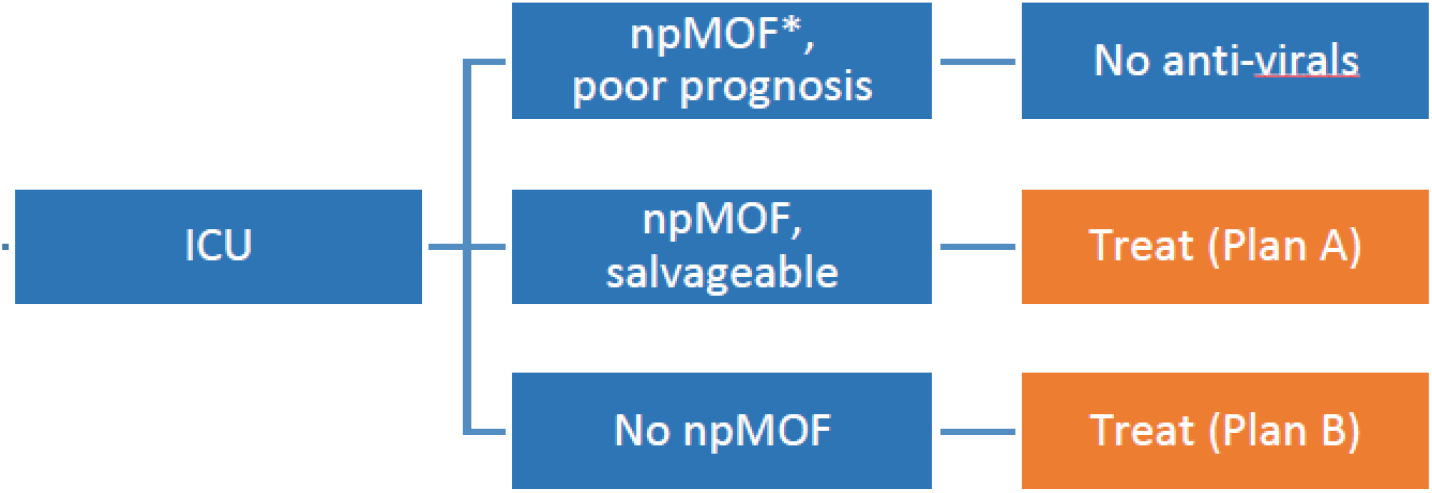
Proposed algorithm for COVID-19 directed medication therapy at RGH. *npMOF = non-pulmonary multiorgan failure

**Figure 3b:**
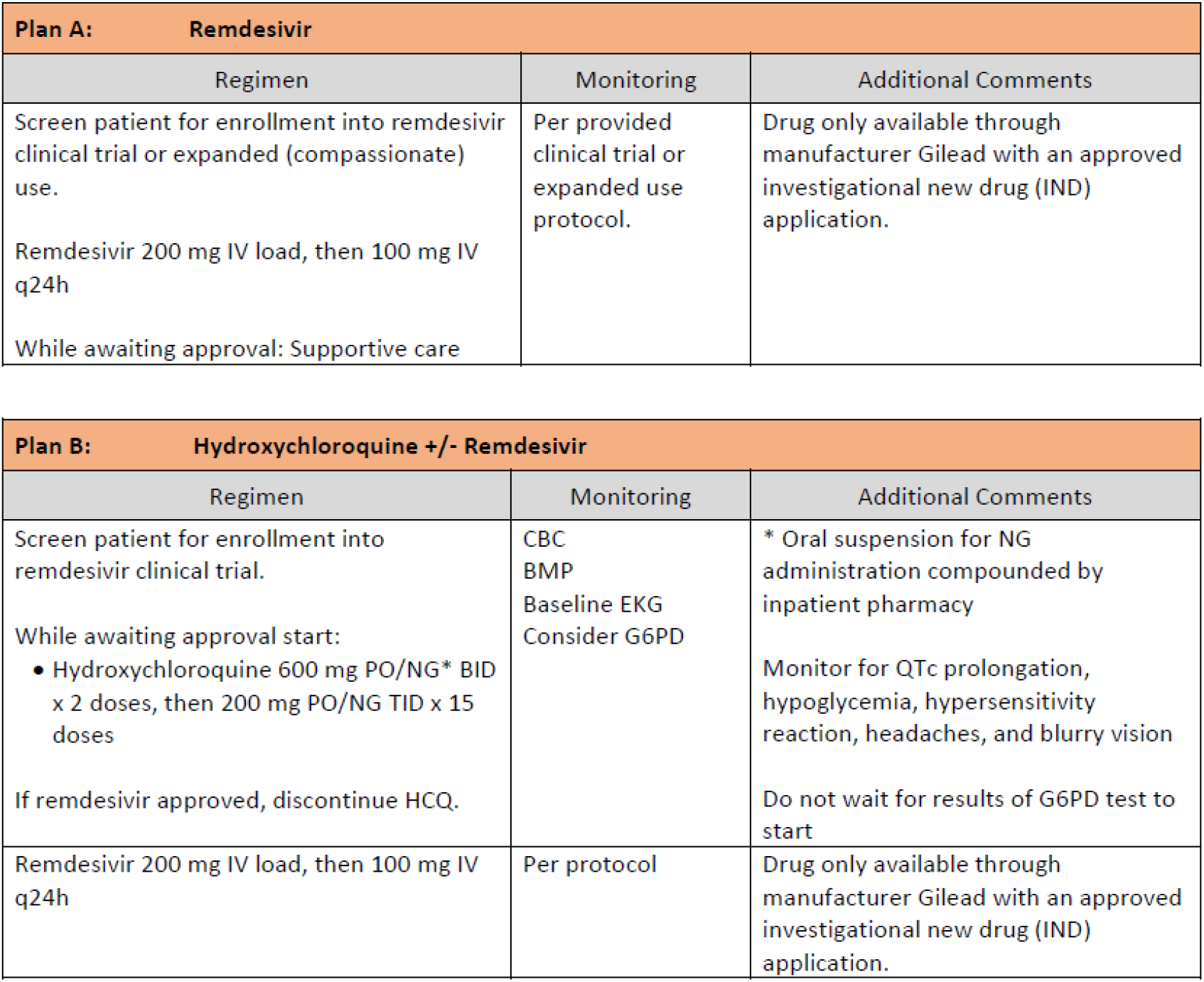
Treatment Plans A and B described

At 15.6%, our case fatality rate (CFR) appears to be lower than prior published literature internationally from Italy, and locally from NYC and Seattle.^21, 22, 5^ This could be due to the fact that our hospital was not yet over-capacity, and we had received timely information and guidance about the natural course and complications of the disease. This suggests that with adequate availability of health care resources, critically ill COVID-19 patients may experience lower morbidity and mortality than suggested by current data.^22^ This re-enforces the concept of ‘flattening the curve’ to prevent strain on the healthcare system.

In our case series, there were no statistically significant differences in patients aged 60 years or older vs. those younger than 60 years of age, which suggests that other prognostic factors, apart from age, play a role in morbidity and mortality in COVID-19. Longer duration of ICU and hospital stays were noted in patients who received IMV. This appears intuitive as patients who undergo IMV usually have more severe disease, more comorbidities and are at higher risk of developing complications during hospital stay (secondary infections, VTE) etc. These numbers need to be interpreted with caution, since they may be an underestimate for the 16 patients that remained in the hospital at time of data calculation.

Regarding complications, VTE events were the most common (15.6%) and additional non-pulmonary organ failure rate was low. Thrombotic complications in patients with severe COVID-19 is an area of debate, with some studies reporting rates of thrombotic complication as high as 31% despite adequate thromboprophylaxis.^23^ The increased risk in thrombotic complications with COVID-19 is difficult to ascertain especially in critically ill patients where risk of clotting is ∼20%.^24.^ It is hypothesized that with severe inflammatory state, there is increase in pro-inflammatory cytokines IL-1, IL-6, and TNF-α, leading to increased thrombin generation and stimulation of the coagulation pathway. Additionally, in hypoxia, there is upregulation of hypoxia-inducible transcription factor (HITF) which stimulates tissue factor and plasminogen activator inhibitor 1 (PAI-1) gene expression, predisposing to increased VTE complications.^25^. Regardless of etiology, the importance of weight-adjusted, renal function based thromboprophylaxis is underscored, as per latest International Society of Thrombosis Hemostasis (ISTH) consensus^26^. To preemptively treat these patients with full dose of anticoagulation seems pre-mature, without stable epidemiological data, and efficacy and safety outcomes from randomized trials.

## Limitations

The main limitation of our study is the small sample size, however, our study focuses on the critically ill patients with most severe disease. Second, due to short follow up, outcomes of the patients that remain in hospital are not known, however we aimed to report the in-hospital outcomes and complications of these patients. Third, patients who had do-not-resuscitate (DNR) orders were included in the NIV group; these patients are likely to be older and sicker, and have higher likelihood of having worse clinical outcomes and complications. Nevertheless, the NIV group only had 1 death compared to 4 deaths in the IMV group. Fourth, we did not include the patients managed in the step-down unit of the hospital, but this came to our advantage as it served as an unbiased measure in selecting the sickest patients in the cohort who were then managed in the ICU. Finally being a retrospective, observational study, there are inherent biases including selection bias, confounding bias and the inability to attribute causation.

## Conclusion

Our case series describes early experience of critically ill patients at a tertiary center upstate NY. Our findings underscore the higher risk of severe illness in older persons and those with pre-existing medical conditions; and also inform us of the prolonged need of critical care resources in those critically ill with COVID-19. These findings can be used to screen patients at higher risk, and to guide resource allocation. Our case fatality rate (CFR) was lower than prior published data, rendering hope that with adequate medical infrastructure and timely resource allocation, mortality from COVID-19 may not be as high as currently reported in parts of the world where healthcare systems have become strained. Nevertheless, much remains unknown about the appropriate ventilation and management strategies of these patients. Large scale prospective studies are needed to further elucidate the efficacy of novel treatments, and identify specific predictors of mortality and other outcomes in the COVID-19 population.

## Data Availability

We collected demographic, clinical, laboratory, radiological, ventilator and treatment data by manual review of electronic medical records (EPIC EMR) of patients admitted to our medical intensive care unit. The data was then de-identified, prior to analysis.

## Acknowledgements

Authors JL and DSS had full access to all of the data in the study and take responsibility for the content of the manuscript, including the data and analysis. All the authors, JL, MSC, NS, BEXT, LTKB, SV, and DSS contributed substantially to the study design, data analysis and interpretation, writing and revision of the manuscript.

## Abbreviations

ACE-i: Angiotensin converting enzyme inhibitor
ARB: Angiotensin receptor blocker
ARDS: Acute Respiratory Distress Syndrome
BiPAP: Bilevel positive airway pressure
CABG: Coronary artery bypass graft
CFR: Case fatality rate
COVID-19: Coronavirus disease 19
CPAP: Continuous positive airway pressure
CRP: C – Reactive Protein
CT: Computed tomography
DVT: Deep vein thrombosis
ECMO: Extra Corporeal Membrane Oxygenation
ESICM: European Society of Intensive Care Medicine
FiO2: Fraction of inspired O2
HFNC: High Flow Nasal Cannula
HITF: Hypoxia-Inducible Transcription Factor
IBM: International Business Machines
ICU: Intensive Care Unit
IL: Interleukin
IMV: Invasive Mechanical Ventilation
IQR: Interquartile Range
ISTH: International Society of Thrombosis Hemostasis
NIV: Non Invasive Ventilation
NY: New York
PAI: Plasminogen activator inhibitor
PaO2: partial pressure of arterial oxygen
PCV: Pressure Control Ventilation
PEEP: Positive End Expiratory Pressure
RGH: Rochester General Hospital
RRH: Rochester Regional Health
RT-PCR: Reverse transcriptase polymerase chain reaction
RSV: Respiratory Syncytial virus
SARS-CoV-2: Severe Acute Respiratory Syndrome Coronavirus 2
SD: Standard Deviation
STEMI: ST segment elevation myocardial infarction
TNF: Tumor necrosis factor
USA: United States of America
VTE: Venous thromboembolism

